# Machine Learning-Supported VTE Risk Assessment using Routinely Collected Electronic Health Record Data

**DOI:** 10.64898/2026.08.18.26360687

**Authors:** Zhuoyu Li, Ekin Yagis, Aya Riad, Oscar Windrath-Carr, Maite Arribas, Toheeb Sodiq, Kathleen Goldsmith, Ben Glampson, Kelsey Flott, Gulammehdi Haji, Zubeyr Khan, Chris Baker, Erik K Mayer

**Affiliations:** Imperial Clinical Analytics, Research and Evaluation (iCARE), NIHR Imperial Biomedical Research Centre, Imperial College Healthcare NHS Trust, London, UK; Faculty of Medicine, Department of Surgery & Cancer, Imperial College London, London, UK

**Keywords:** Venous thromboembolism, Electronic health records, Risk assessment, Machine learning, Clinical decision support

## Abstract

**Objective:** Venous thromboembolism (VTE) is a leading cause of preventable inpatient mortality, while the real-world performance of mandated risk assessment and the potential for automating using electronic health record (EHR) data remain unclear. We evaluated completion, concordance with structured EHR data, clinical validity, and feasibility of EHR-supported VTE risk assessment assisted by machine learning.

**Materials and Methods:** We analysed 577,904 admissions and 726,896 VTE assessment forms across five NHS hospitals between 2015 and 2025. Concordance and clinical validity were evaluated in 87,118 completed assessments from 2023 to 2024. Machine learning models compared clinician-recorded and EHR-derived risk factors in 74,372 encounters with an assessment completed within 14 hours of admission.

**Results:** Overall completion was 96.7%, and timely completion improved from 47.4% in 2015 to 90.5% in 2024. Agreement between forms and EHR data was good for common risk factors (e.g. age ≥ 60 years: precision 0.95, recall 0.77), but low-prevalence variables were often under-documented in the forms (e.g. critical care admission: precision 0.20, recall 0.14). Despite these discrepancies, form-derived thrombosis risk was associated with increased VTE incidence (adjusted OR 3.31, 95% CI 2.81–3.90). Models using EHR-derived factors available within 14 hours achieved an AUROC of 0.709, compared with 0.704 for clinician-recorded variables.

**Discussion:** Much of the information the risk assessment requires already exists in structured EHR data, and EHR-derived variables contained discriminative information comparable to that in clinician-recorded factors.

**Conclusion:** Routinely collected EHR data could support real-time pre-population of structured VTE assessments, thereby reducing documentation burden while preserving clinical validity.

## Background and Significance

Venous thromboembolism (VTE) refers to the formation of blood clots in a vein, including deep vein thrombosis (DVT) and pulmonary embolism (PE). VTE is one of the leading causes of preventable death and long-term disability [1]. Hospitalisation, particularly for acute medical illness or surgery, substantially increases the risk of blood clots due to reduced mobility, inflammation, and physiological stress [2–4].

The National Institute for Health and Care Excellence (NICE) guideline NG89 mandates that all adult patients aged 16 years or over admitted to an NHS hospital undergo a formal, structured VTE risk assessment at or near the time of admission. The guideline further specifies that thromboprophylaxis should be initiated as soon as possible and within 14 hours of admission. Reassessment is required whenever the patient’s clinical condition changes [5]. Beyond the UK NHS setting, structured risk assessment models, including Caprini, Padua, and IMPROVE, are widely used to support VTE prevention across different patient settings [6–8].

Despite widespread implementation of these tools, evidence suggests that translating policy into consistent clinical practice remains challenging [9,10]. Risk assessment is typically performed manually within electronic health record (EHR) systems, requiring clinicians to re-enter information that is often already available in structured form elsewhere in the EHR. This duplication consumes clinician time and introduces transcription error, inconsistency, and the risk that high-risk patients are misclassified or overlooked. Prior studies have documented high rates of incomplete documentation, under-reporting of low-prevalence risk factors, and inconsistency between repeated assessments for the same patient [11,12].

Structured EHR data and machine learning (ML) have stimulated interest in automated and semi-automated clinical risk assessment [13]. In the context of VTE, studies have explored EHR-driven VTE risk prediction using logistic regression, tree-based models, and more recently deep learning architectures across diverse settings [14–16]. A large multi-cohort study demonstrated that gradient boosting models trained on structured EHR data can outperform the conventional clinical risk score Padua for both VTE diagnosis prediction and 1-year risk prediction [17].

Beyond prediction, recent studies indicate that EHR-integrated clinical decision support (CDS) can improve VTE prophylaxis practice and reduce preventable harm. Mandatory CDS tool and computer alert systems have increased appropriate thromboprophylaxis rates and reduced VTE incidence in hospitalised patients [18,19]. The ongoing VTE-AI trial further reflects growing interest in real-time ML-driven EHR interventions [20]. EHR-derived data may also support automated or semi-automated completion of structured risk assessment fields through form pre-population before clinician review, potentially reducing manual data entry, lessening clinician burden, and improving completeness and consistency of documentation.

However, important barriers remain to the use of these approaches in routine care. Many existing models are designed primarily to predict VTE outcomes rather than to support structured risk assessment for real-time clinical use. Information available only later during admission, or on variables not consistently accessible at the point of care, may limit clinical applicability. Whether routinely collected EHR data can reproduce clinician-completed structured risk assessments at the level of individual risk factors within a nationally mandated risk assessment framework remains an important unassessed question.

We address this question using a large-scale, multi-hospital dataset from five hospitals within Imperial College Healthcare NHS Trust in North-West London, spanning a decade of routinely collected EHR data and over two million hospital encounters between January 2015 and January 2025. We evaluate (i) completeness and timeliness of VTE risk assessment, (ii) agreement between clinician-recorded and EHR-derived risk factors, (iii) associations between risk assessment results, prophylaxis prescribing, and VTE outcomes, and (iv) the feasibility of automated pre-population of structured VTE risk assessment forms using data available at the point of care by developing and comparing ML models. By examining whether re-using information already recorded in the EHR can support VTE risk assessment and prevention workflows, this study aims to inform the development of embedding scalable, data-driven risk stratification within routine NHS workflows, paving the way for more integrated clinical decision support systems in clinical practice.

## Materials and methods

### Study cohort

A retrospective study was conducted across five secondary care hospitals within Imperial College Healthcare NHS Trust in North-West London. The cohort comprised all inpatients aged ≥16 years, excluding those in the maternity period by specialty coding, admitted between 31 January 2015 and 31 January 2025. Admissions were included if they required completion of a VTE risk assessment according to local clinical policy. Detailed exclusions are provided in Supplementary Note S1. All patient data were anonymised, accessed and analysed within the Imperial Secure Data Environment (SDE).

### Ethics

This study was undertaken within a research database that was given favourable ethics approval by the South West - Central Bristol Research Ethics Committee (reference 21/SW/0120; IRAS project ID 282093). As only anonymised data were used, individual informed consent was not required.

### VTE risk assessment structure

The VTE risk assessment is a three-stage workflow embedded within the electronic health record system (Figure 1). The initial stage is mobility screening, which determines whether further assessment is required. Patients identified as surgical, or as medical patients expected to have reduced mobility, proceed to stage two, structured evaluation of thrombosis and bleeding risks. These risk factors consist of four sections: patient- and admission-related thrombosis and bleeding risks. Clinicians select applicable risk factors or ‘None’. The presence of any corresponding factor classifies the patient as having thrombosis or bleeding risk. Patients with thrombosis risk proceed to stage three, a mandatory prescribing screen in which clinicians make a thromboprophylaxis decision and, if not prescribing, provide a free-text reason. If a patient has bleeding risk, clinicians needed to balance the thrombosis and bleeding risks.

**Figure 1.**
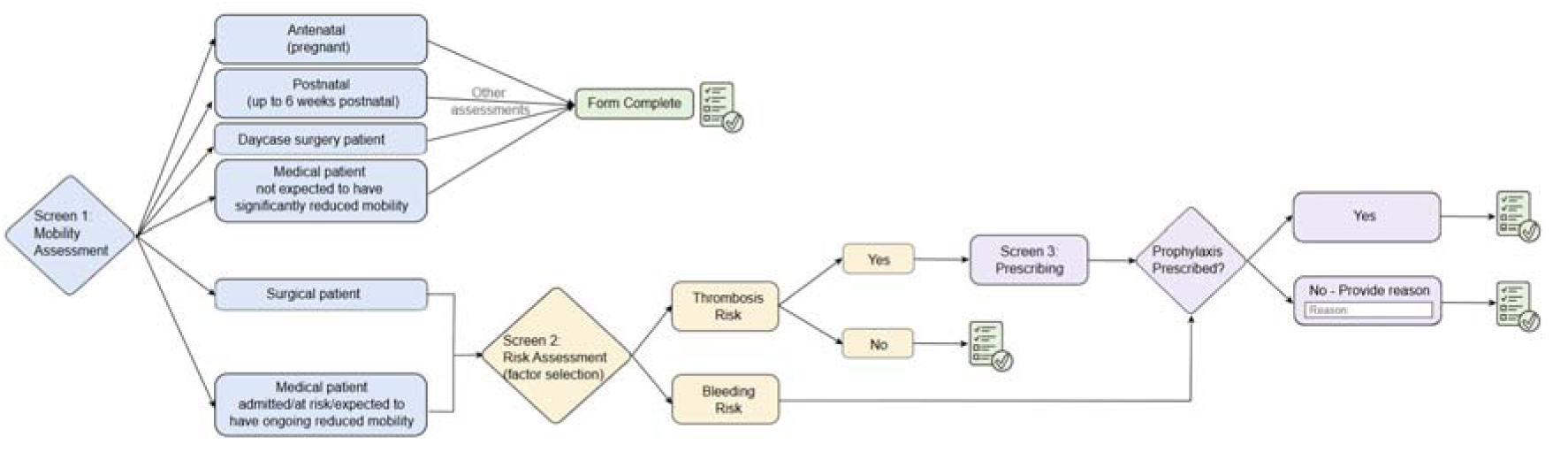
Workflow of the VTE risk assessment tool and prescribing pathway. Alt text: Flow diagram of the three-stage VTE assessment. Mobility screening proceeds surgical patients and medical patients expected to have reduced mobility to risk-factor selection. Patients with thrombosis risk proceed to a prophylaxis decision with consideration of bleeding risk. A reason is required when prophylaxis is not prescribed.

While the overall assessment pathway remained consistent throughout the study period, three form versions were used: before 22 March 2017, 22 March 2017 to 17 October 2022, from 17 October 2022. Items were grouped if wording differences were minimal and reflected the same underlying clinical meaning.

### Data definitions

Demographic variables, including age, sex at birth, ethnicity, and Index of Multiple Deprivation (IMD), were obtained from structured electronic health records. For descriptive reporting, ethnicity was grouped as White, Black, Asian, Mixed, Other ethnicity, or Missing/Unknown. IMD deciles range from 1 (the most deprived 10% of areas in England) to 10 (the least deprived 10%). Ethnicity and IMD described the cohort, and the primary multivariable models were adjusted for age and sex.

VTE events, including DVT and PE, were defined using ICD-10 and SNOMED codes. Risk factors were mapped to ICD-10 diagnosis codes, SNOMED clinical terminology, and OPCS-4 procedural codes from routinely collected EHR. The mapping used NICE Guidelines [5] and Health Data Research UK’s (HDR UK) Phenotype Library [21] where available and was confirmed by two clinicians. Each risk factor was assigned a clinically appropriate temporal window, distinguishing between pre-existing conditions and those arising during the admission. Pre-existing conditions, such as known thrombophilia, were defined using all structured history prior to admission. Admission-related variables were restricted to records or observations during the encounter. Full definitions and model-specific temporal windows are provided in Supplementary Table S1.

Thromboprophylaxis includes mechanical and pharmacological prophylaxis. Mechanical prophylaxis included anti-embolism stockings and intermittent pneumatic compression (IPC), as recorded in structured procedures. Pharmacological prophylaxis was defined as the prescription of a prophylactic indication, dosage, and frequency consistent with standard rules for low molecular weight heparin (LMWH), unfractionated heparin (UFH), or direct oral anticoagulant (DOAC). Detailed medication definitions, including drug names, dosing, frequency, and prescribing criteria, are provided in Supplementary Table S2.

### Completion and timeliness

Form completion was defined by verification of a recorded assessment form. Completion was calculated among all recorded forms, overall and by calendar year. Timely completion was evaluated within 14 hours of hospital admission, in accordance with NICE guidance. Time to completion was calculated as the interval between admission and the first recorded VTE risk assessment within the same encounter. Admissions without a recorded assessment were classified as not timely. Analyses were stratified by mobility group, and annual group distributions were examined.

### Concordance between clinical-recorded and EHR-derived risk factors

To evaluate concordance, analyses were restricted to 1 January 2023 to 31 December 2024, during which a single form version was in use, thereby minimising heterogeneity arising from form revisions. We further focused on surgical patients and medical patients with expected reduced mobility, as these groups require full-stage risk assessments. Forms with missing values in any required field or that were not completed (i.e. not verified) were excluded (Supplementary Note S2). For each risk factor, clinician-recorded selections were compared with corresponding factors independently derived from EHR data using predefined algorithms. Agreement was quantified using precision, recall, and specificity. Aggregated thrombosis and bleeding classifications were derived from EHR data using the same logic as the form. Clinician-recorded prophylaxis decisions were compared with the presence of corresponding prophylaxis identified from the EHR prescribing table within the same encounter, excluding home medications and discharge prescriptions.

### Statistical analyses

To evaluate the clinical validity of the VTE risk assessment and its relationship with clinical decision-making and patient outcomes, three key questions were addressed: (i) whether patients identified as having thrombosis risk had higher rates of VTE events than those without; (ii) whether patients identified as having bleeding risk received pharmacological prophylaxis less frequently than those without; and (iii) whether documented prophylaxis was associated with reduced VTE event rates among patients with identified thrombosis risk.

Associations between categorical variables were assessed using chi-squared tests, and effect sizes were quantified using odds ratios with corresponding 95% confidence intervals (CIs). Multivariable logistic regression specified VTE occurrence as the outcome, with thrombosis risk, bleeding risk, and prophylaxis as predictors, adjusted for age and sex. Sensitivity analyses substituted EHR-derived definitions for risk factors and for prophylaxis separately and jointly. P<0.001 was considered statistically significant.

### Machine learning analyses

Proof-of-concept classifiers compared EHR-derived variables with clinician-recorded binary thrombosis risk factors recorded during the same encounter. For comparability, EHR-derived features were binarised to match the form. No data-driven feature selection was applied to Models C, E1 and E2, as the objective was not to optimise predictive performance but to evaluate whether EHR-derived variables could reproduce the predictive value of the clinician-recorded risk factors. Analyses were conducted on a further restricted assessment subset, comprising the first completed VTE risk assessment forms within 14 hours of admission. The outcome was any VTE event recorded during the same hospital encounter.

Four feature configurations were constructed to reflect different levels of information availability and extraction across the clinical workflow (Figure 2). Model-C (clinician model) was a baseline using structured clinician-recorded risk factors directly from the form. Model-E1 (full EHR model) used all EHR-derived risk factors. Model-E2 (time-restricted EHR model) included only variables available within 14 hours of admission. This ensured temporal alignment between realistically accessible information and the timing of risk assessment in clinical practice. Model-E3 (reduced-feature EHR model) evaluated whether a simplified feature set could maintain classification performance while improving interpretability and potential deployability. Features were selected based on Model-E1 SHAP (SHapley Additive exPlanations) values and clinician-recorded selection frequency. The full list of included features, their definitions, and temporal windows is provided in Supplementary Table S1.

**Figure 2.**
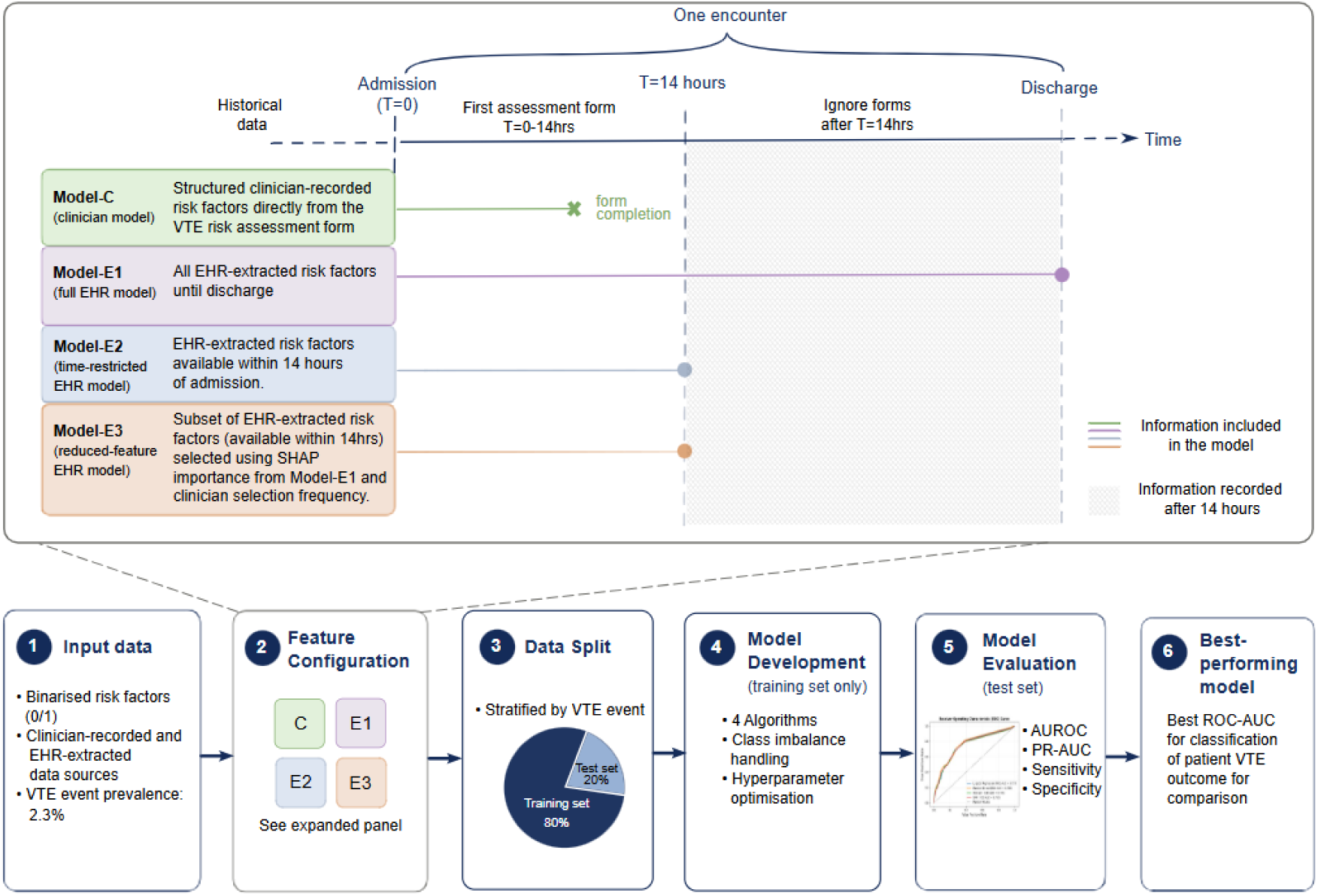
Feature configuration and machine learning pipeline for automated VTE risk assessment. a) The temporal structure of a hospital encounter and the feature availability windows used to define four modelling configurations. b) The machine learning pipeline. Alt text: Two-panel diagram. The first panel maps four feature configurations to the admission timeline: clinician-recorded form data, all EHR data through discharge, EHR data within 14 hours, and a reduced subset of 14-hour features. The second shows the modelling pipeline from binary inputs through feature configuration, stratified data splitting, model development, evaluation, and best-model selection.

Each feature configuration was then evaluated using the same supervised machine learning pipeline. Four supervised classification algorithms were evaluated: logistic regression, random forest, XGBoost, and support vector machine (SVM). Data were split into training (80%) and testing (20%) sets at patient level and stratified by VTE prevalence. All encounters from the same patient were assigned to one set. Hyperparameters were optimised using grid search with five-fold cross-validation and folds grouped by patient, and all preprocessing steps were performed within training folds. For each feature configuration, the classifiers were selected by mean cross-validated AUROC. Given the low prevalence of VTE (2.3% of encounters), class imbalance was addressed using class weighting as the primary approach, applied only within the training data. Undersampling and SMOTE (Synthetic Minority Over-sampling Technique), were evaluated as sensitivity analyses using the imbalanced-learn library.

Model performance was assessed using multiple metrics. The area under the receiver operating characteristic curve (AUROC) represented the trade-off between true positive and false positive rates across thresholds. The area under the precision-recall curve (PR-AUC) summarised the balance between correctly identifying VTE cases while avoiding false positives across all probability thresholds, with random performance equal to event prevalence (0.023). Additional metrics included recall and specificity. The metrics’ 95% CIs were calculated from the 2.5th and 97.5th percentiles of 1,000 patient-level bootstrap resamples of the test set. SHAP values were computed on the held-out test set.

Analyses used Python (3.10.11), scikit-learn (1.5.1), XGBoost (1.5.2), imbalanced-learn (0.12.4), and SHAP (0.44.0).

## Results

### Cohort summary

A total of 2,105,339 hospital encounters were identified between 31 January 2015 and 31 January 2025. After applying eligibility criteria, 577,904 inpatient admissions with 726,896 VTE risk assessment forms recorded were included in the overall analytical cohort.

Concordance and clinical validity analyses used 87,118 eligible and completed assessments from 85,325 encounters in 2023-2024. For machine learning analyses, the cohort was further restricted to 74,372 assessments representing the earliest completed assessments within 14 hours of admission per encounter. Cohort selection is shown in Figure 3 and characteristics in Table 1.

**Figure 3.**
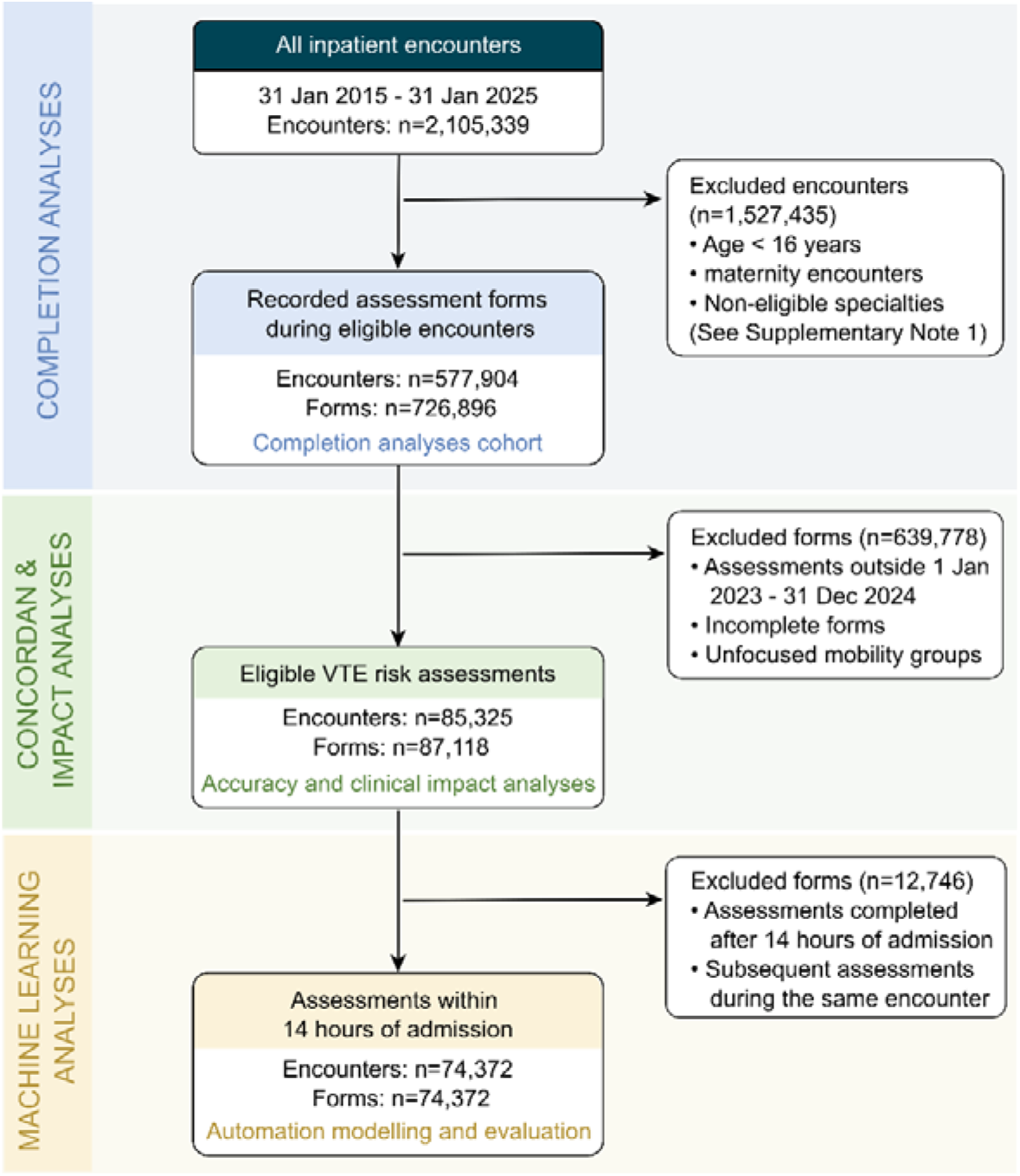
Study cohort selection and derivation of assessment subsets for completion, concordance, clinical impact, and machine learning analyses of VTE risk assessments. In the exclusion box, maternity encounters are defined by admission treatment-function or specialty coding. VTE, venous thromboembolism. Alt text: Cohort flow diagram. Of 2,105,339 encounters screened, 577,904 eligible encounters with 726,896 forms entered completion analyses; 85,325 encounters with 87,118 forms entered concordance and impact analyses; and 74,372 encounters with assessments completed within 14 hours entered machine learning analyses. Exclusion reasons are shown at each stage.

**Table 1.**
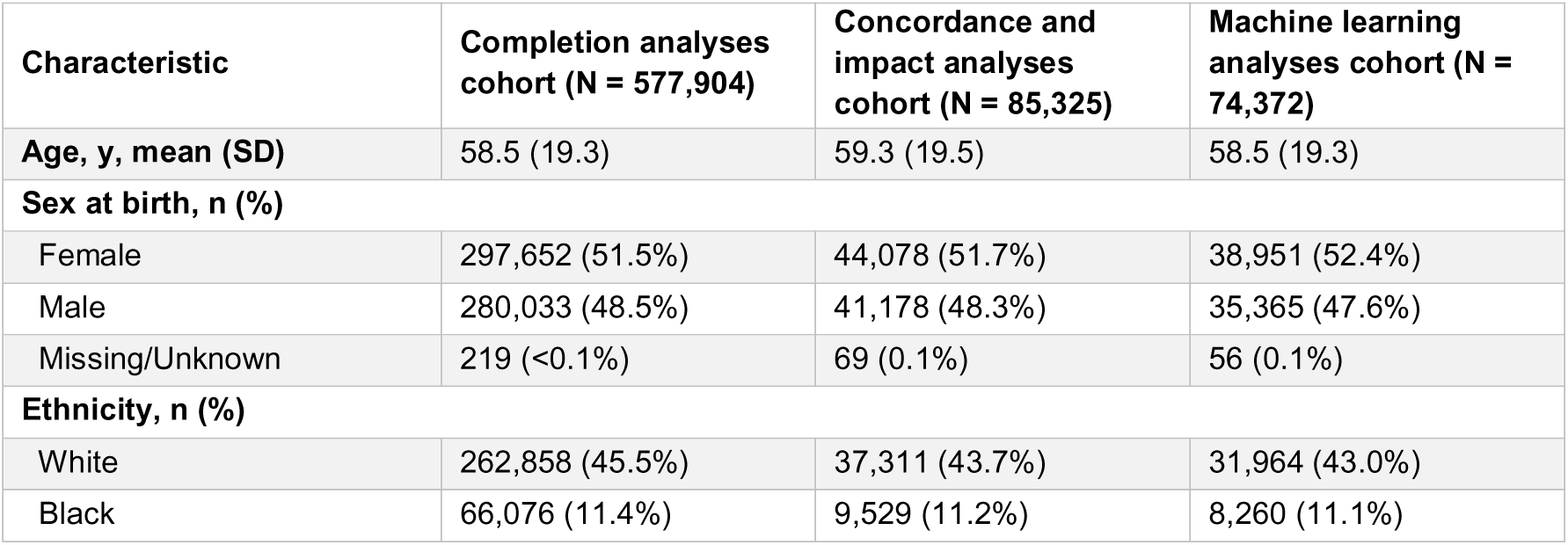
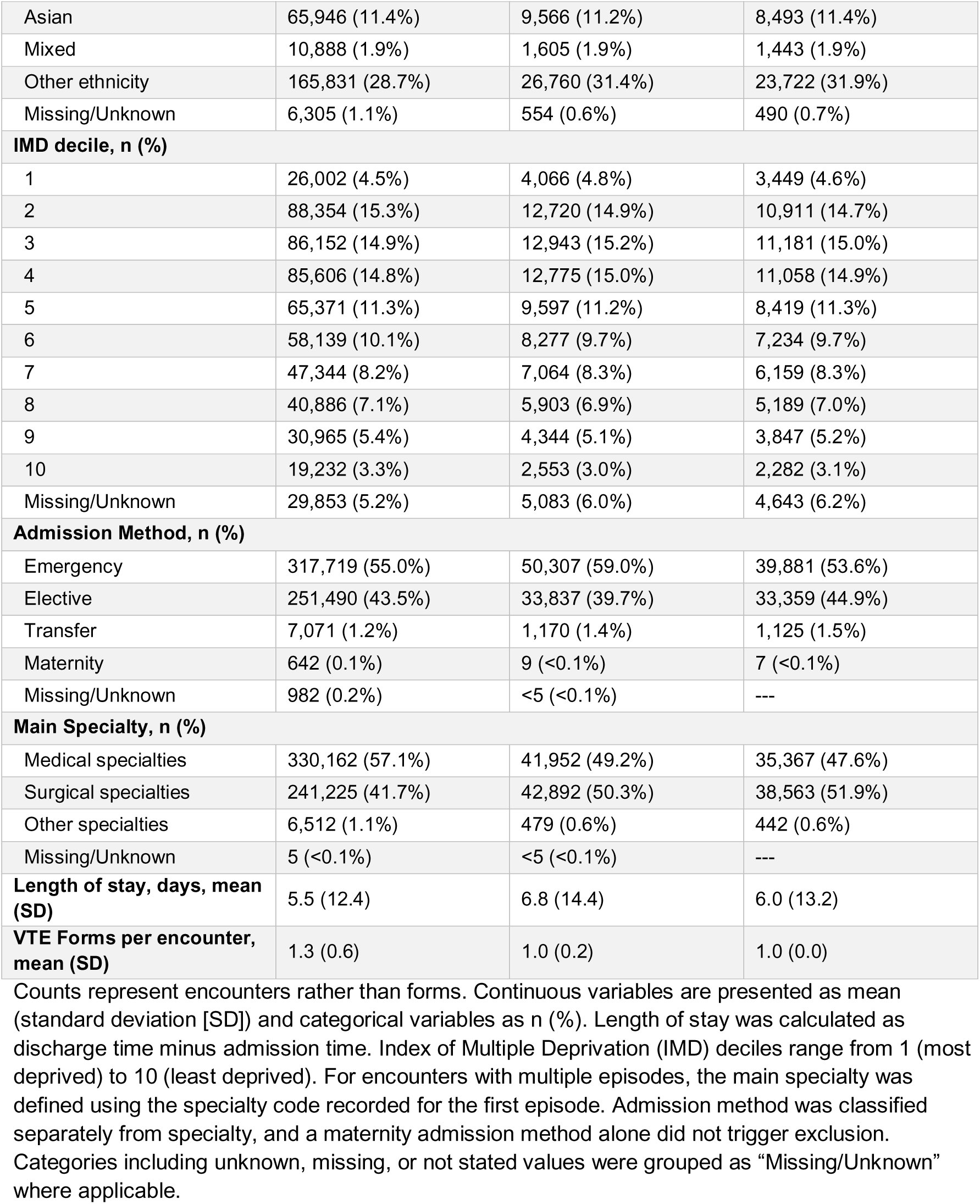
Cohort characteristics across the completion, concordance and impact, and machine learning analyses cohorts.

### Completion rates

Among all 726,896 recorded VTE risk assessments, 702,627 were completed, corresponding to an overall completion rate of 96.66%. Completion rates varied over time, with the highest observed in 2015 at 99.66% and the lowest in 2020 at 93.20%, followed by a gradual recovery to 98.58% in January 2025 (Figure 4a). Despite high overall completion, the proportion of assessments completed within 14 hours was lower, at 83.90%, increasing from 47.35% in 2015 to 90.50% in 2024.

**Figure 4.**
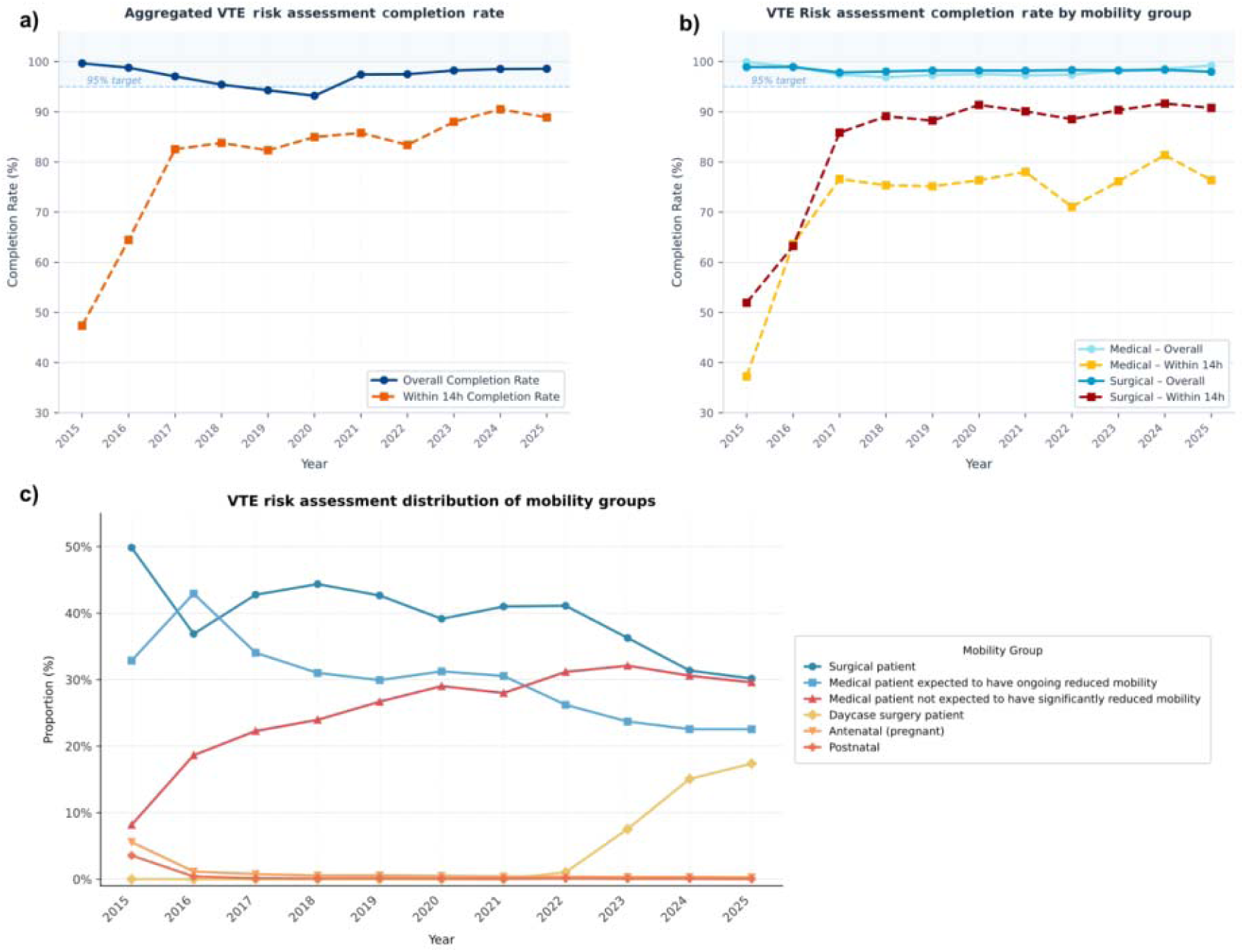
Temporal trends in VTE risk assessment completion, timely completion, and mobility-group composition. a). Overall completion rate (blue solid line) and completion rate within 14 hours of admission (orange dashed line). b) Overall completion rate (solid lines) and timely completion rate (dashed lines) stratified by mobility group, including surgical patients (“Surgical”) and medical patients expected to have ongoing reduced mobility (“Medical”). c) Annual distribution of mobility groups (proportions) among all admissions at the initial screening stage. Data for 2025 include admissions through 31 January 2025 only and therefore do not represent a complete calendar year. Alt text: Three line charts showing annual form completion, completion within 14 hours, and mobility-group composition. Overall completion remains high; timely completion improves, with surgical assessments more timely than reduced-mobility medical assessments. The admission composition changes over time.

Among all assessments, 16,072 (2.21%) had invalid or missing mobility status. The remaining assessments were classified as 191,019 (26.28%) medical patients not expected to have reduced mobility, 21,132 (2.91%) day-case surgical patients, 4,047 (0.56%) antenatal patients, and 1,080 (0.15%) postnatal patients within six weeks of delivery. The remaining assessments consisted of 211,376 (29.08%) medical patients expected to have reduced mobility and 282,170 (38.82%) surgical patients, who proceeded to second-stage assessment, namely risk factor selection.

Timely completion differed between groups (Figure 4b). Surgical patients showed higher rates of timely assessment, reaching approximately 90% from 2018 onwards, whereas medical patients demonstrated lower rates throughout the study period, improving from 37.20% in 2015 to 76.56% in 2025. The proportions of surgical and reduced-mobility medical admissions declined, while low-risk medical and day-case surgical groups increased (Figure 4c). Data for 2025 include January only.

### Concordance of form and EHR data

Of 92,710 eligible assessments in 2023-2024, 5,592 were excluded due to missing values in any required fields or incomplete forms (details in Supplementary Note S2), leaving 87,118 assessments across 85,325 encounters. Within these forms, 20,743 (23.81%) had no recorded risk factors across all four sections (i.e. clinicians selected four ‘none’s), while 66,375 (76.19%) selected at least one risk factor.

At the individual risk factor level, agreement varied substantially across both patient-related and admission-related variables (Table 2). Frequently selected structured risk factors, such as *age* ≥*60 years old*, showed relatively high agreement, with precision of 0.95 and recall of 0.77. In contrast, less frequently selected factors, such as *critical care admission* and *use of hormone replacement therapy*, showed low precision (0.20 and 0.01, respectively) and recall (0.14 and 0.02, respectively). Several risk factors exhibited notably low recall despite moderate to high specificity, indicating that a substantial proportion of cases identified from the EHR were not captured in the assessment form. For example, *personal or first-degree relative history with VTE* showed a specificity of 0.99 and a recall of only 0.05.

**Table 2.** Comparison of clinician-recorded and EHR-derived extraction of VTE risk factors.

| Section | Risk factor | Clinician-recorded n (%) | EHR-derived n (%) | Precision | Recall | Specificity |
| --- | --- | --- | --- | --- | --- | --- |
| **Thrombosis risk – patient related** | Age equal to or more than 60 years | 36901 (42.36%) | 45680 (52.43%) | 0.95 | 0.77 | 0.95 |
|  | Obesity BMI > 30 kg/m<sup>2</sup> | 7467 (8.57%) | 12138 (13.93%) | 0.44 | 0.27 | 0.94 |
|  | Use of hormone replacement therapy | 222 (0.25%) | 165 (0.19%) | 0.01 | 0.02 | 1.00 |
|  | One or more significant medical comorbidities | 23503 (26.98%) | 38256 (43.91%) | 0.61 | 0.37 | 0.81 |
|  | Varicose veins with phlebitis | 670 (0.77%) | 141 (0.16%) | 0.01 | 0.05 | 0.99 |
|  | Personal history or first-degree relative with a history of VTE | 1262 (1.45%) | 14829 (17.02%) | 0.61 | 0.05 | 0.99 |
|  | Active cancer or cancer treatment | 8026 (9.21%) | 14244 (16.35%) | 0.83 | 0.47 | 0.98 |
|  | Dehydration | 7107 (8.16%) | 1976 (2.27%) | 0.07 | 0.24 | 0.92 |
|  | Pregnancy or <6 weeks postpartum | 491 (0.56%) | 9 (0.01%) | 0.00 | 0.11 | 0.99 |
|  | Use of oestrogen-containing contraceptive therapy | 116 (0.13%) | 12 (0.01%) | 0.01 | 0.08 | 1.00 |
|  | Known thrombophilia | 247 (0.28%) | 278 (0.32%) | 0.17 | 0.15 | 1.00 |
|  | None | 31350 (35.99%) | 22419 (25.73%) | 0.56 | 0.78 | 0.79 |
| **Thrombosis risk – admission related** | Hip or knee replacement | 1175 (1.35%) | 913 (1.05%) | 0.61 | 0.78 | 0.99 |
|  | Hip fracture | 670 (0.77%) | 579 (0.66%) | 0.48 | 0.56 | 1.00 |
|  | Total anaesthetic time + surgical time >90 minutes | 6720 (7.71%) | 1669 (1.92%) | 0.06 | 0.23 | 0.93 |
|  | Critical care admission | 755 (0.87%) | 1051 (1.21%) | 0.20 | 0.14 | 0.99 |
|  | Surgery with significant reduction in mobility | 1739 (2.00%) | 1175 (1.35%) | 0.09 | 0.14 | 0.98 |
|  | Significantly reduced mobility for 3 days or more | 17282 (19.84%) | 10780 (12.37%) | 0.22 | 0.35 | 0.82 |
|  | Other thrombosis risk - specify in notes | 817 (0.94%) | 559 (0.64%) | 0.01 | 0.01 | 0.99 |
|  | Acute surgical admission with inflammatory or intra-abdominal condition | 2289 (2.63%) | 4605 (5.29%) | 0.18 | 0.09 | 0.98 |
|  | Surgery involving pelvis or lower limb and total anaesthetic time > 60 minutes | 4131 (4.74%) | 9249 (10.62%) | 0.16 | 0.07 | 0.96 |
|  | None | 57417 (65.91%) | 63376 (72.75%) | 0.78 | 0.71 | 0.48 |
| **Bleeding risk – patient related** | Acute Stroke | 2190 (2.51%) | 442 (0.51%) | 0.11 | 0.54 | 0.98 |
|  | Acquired bleeding disorders e.g. acute liver failure | 53 (0.06%) | 3647 (4.19%) | 0.15 | 0.00 | 1.00 |
|  | Untreated inherited bleeding disorders | 7 (0.01%) | 148 (0.17%) | 0.57 | 0.03 | 1.00 |
|  | Concurrent use of other anticoagulants | 3559 (4.09%) | 13109 (15.05%) | 0.32 | 0.09 | 0.97 |
|  | Active bleeding or risk of bleeding | 7446 (8.55%) | 13570 (15.58%) | 0.38 | 0.21 | 0.94 |
|  | Uncontrolled Hypertension (> 230/120 mmHg) | 80 (0.09%) | 49 (0.06%) | 0.11 | 0.18 | 1.00 |
|  | None | 73755 (84.66%) | 60579 (69.54%) | 0.72 | 0.88 | 0.23 |
| **Bleeding risk – admission related** | Lumbar Puncture /epidural/spinal anaesthesia within the previous 4 hours | 792 (0.91%) | 68 (0.08%) | 0.01 | 0.07 | 0.99 |
|  | Neurosurgery, spinal surgery or eye surgery | 3468 (3.98%) | 3924 (4.50%) | 0.44 | 0.39 | 0.98 |
|  | Lumbar puncture/epidural/spinal anaesthesia expected in next 12 hours | 860 (0.99%) | 182 (0.21%) | 0.05 | 0.23 | 0.99 |
|  | Other procedure with high bleeding risk | 943 (1.08%) | 5563 (6.39%) | 0.09 | 0.02 | 0.99 |
|  | None | 81180 (93.18%) | 77594 (89.07%) | 0.91 | 0.95 | 0.24 |
| **Aggregated level** |  |  |  |  |  |  |
| **VTE risk** |  | --- | --- | 0.89 | 0.82 | 0.64 |
| **Bleeding risk** |  | --- | --- | 0.51 | 0.27 | 0.84 |

At the aggregated level, performance differed between thrombosis risk and bleeding risk identification. For thrombosis risk, agreement between clinician-recorded assessment and EHR-derived classification had a precision of 0.89 and recall of 0.82. For bleeding risk, precision was 0.51 and recall was 0.27. Corresponding confusion matrices are provided in Supplementary Figure S1.

For thromboprophylaxis, 24,075 assessments (27.63%) recorded no thrombosis risk factors and therefore did not proceed to the prophylaxis screening. Among the remaining 63,043 assessments, agreement with EHR prescribing data was 71.71%. This comprised 32,456 (51.48%) assessments with prophylaxis documented in both sources and 12,754 (20.23%) with no prophylaxis in either. Discrepancies were observed in 7,579 (12.02%) assessments where prophylaxis was recorded in the form but not prescribing data, and 10,254 (16.27%) where prophylaxis was prescribed but not recorded in the form.

### Clinical impact

In univariable analyses, thrombosis risk was associated with VTE (OR 4.05, 95% CI 3.45– 4.76; p<0.001). Within the thrombosis-risk group, bleeding risk was associated with lower prophylaxis prescribing (OR 0.42, 95% CI 0.41–0.44; P<0.001). Clinician-recorded prophylaxis was not significantly associated with VTE (OR 0.91, 95% CI 0.83–1.00; P=0.06; Figure 5). Analysis counts are provided in Supplementary Tables S3 and S4.

**Figure 5.**
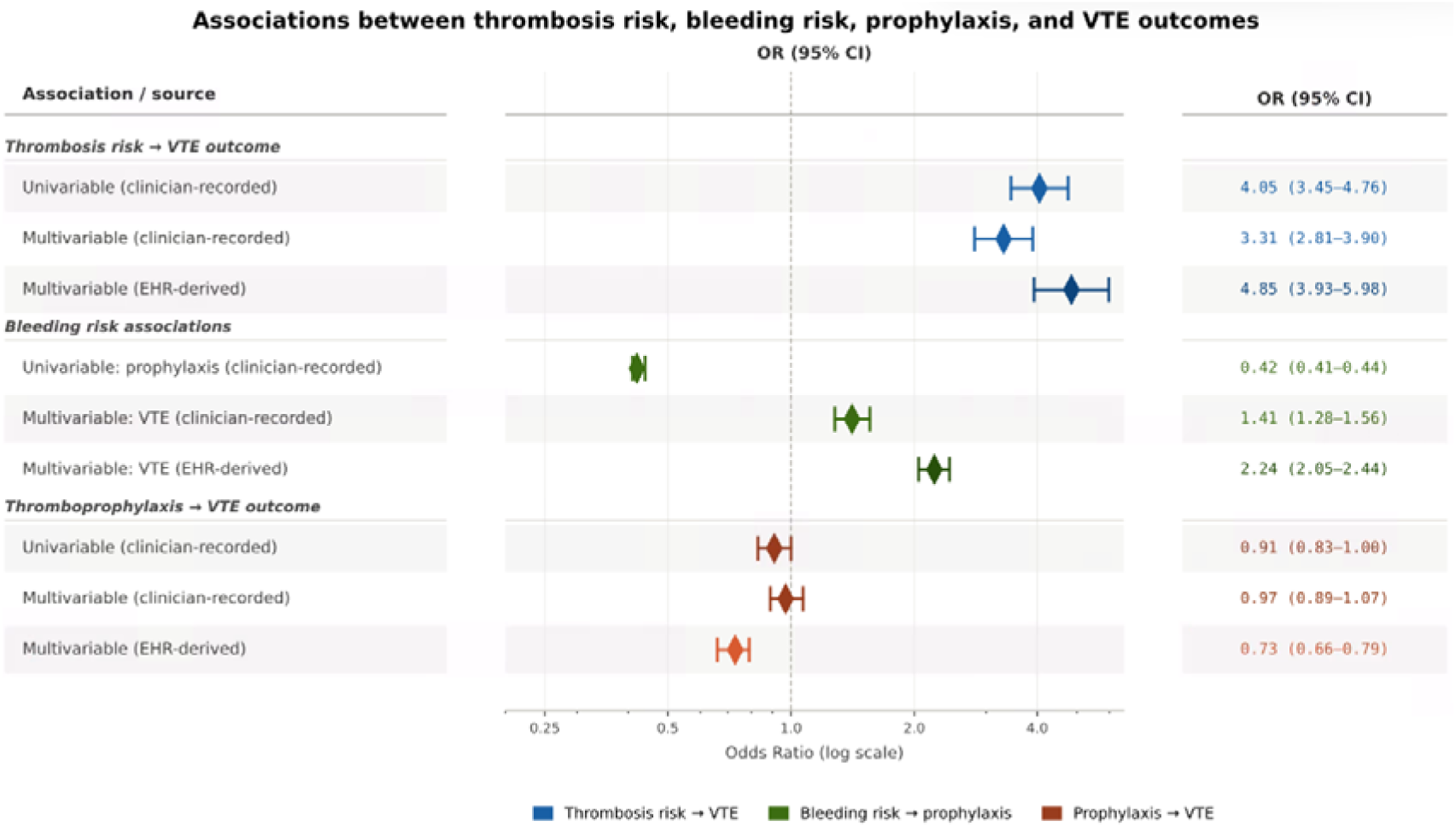
Associations between VTE risk assessment variables, thromboprophylaxis prescribing, and VTE outcomes using clinician-recorded and EHR-derived risk factors. Forest plot showing odds ratios (ORs) and 95% confidence intervals (CIs). The univariable analyses show associations between thrombosis risk and VTE occurrence (blue), bleeding risk and thromboprophylaxis prescribing (green), and thromboprophylaxis and VTE occurrence (orange). The multivariable analyses show the adjusted associations of thrombosis risk (blue), bleeding risk (green), and thromboprophylaxis (orange) with VTE occurrence, using two matched-source models, i.e. both clinician-recorded or EHR-derived definitions, adjusted for age and sex. Sensitivity models are reported in Table 3. Odds ratios are displayed on a logarithmic scale, with the dashed vertical line indicating an OR of 1.0 (no association). EHR, electronic health record; VTE, venous thromboembolism. Alt text: Forest plot of odds ratios and 95% confidence intervals, comparing associations based on clinician-recorded and EHR-derived definitions. Thrombosis and bleeding risk are associated with higher VTE odds in adjusted models. Clinician-recorded prophylaxis has no statistically significant association, whereas EHR-derived prescribing has an inverse association.

**Table 3.** Multivariable logistic regression models for VTE outcomes using clinician-recorded and EHR-derived definitions. Odds ratios (ORs) and 95% confidence intervals (CIs) are shown for models using clinician-recorded (CR) or EHR-derived risk factors and prophylaxis. CR risk refers to clinician-recorded risk factors documented in the assessment form, whereas EHR risk refers to risk factors derived from structured EHR data. CR prophylaxis represents prophylaxis decisions recorded in the assessment form, while EHR prophylaxis represents prophylaxis prescriptions derived from the EHR. Models were adjusted for age and sex.

| Variable | CR risk + CR prophylaxis | CR risk + EHR prophylaxis | EHR risk + CR prophylaxis | EHR risk + EHR prophylaxis |
| --- | --- | --- | --- | --- |
| Thrombosis risk | 3.31 (2.81–3.90) | 3.59 (3.07–4.19) | 4.31 (3.50–5.32) | 4.85 (3.93–5.98) |
| Bleeding risk | 1.41 (1.28–1.56) | 1.39 (1.26–1.53) | 2.18 (2.00–2.38) | 2.24 (2.05–2.44) |
| Prophylaxis prescribing | 0.97 (0.89–1.07) | 0.72 (0.66–0.79) | 1.18 (1.08–1.29) | 0.73 (0.66–0.79) |

In the multivariable VTE-outcome models adjusted for age and sex (Table 3), thrombosis risk remained associated with higher VTE odds across all four models (ORs 3.31-4.85), as did bleeding risk (ORs 1.39-2.24). Clinician-recorded prophylaxis decisions showed no significant association with VTE when paired with clinician-recorded risk (OR 0.97, 95% CI 0.89–1.07), but were associated with higher odds when paired with EHR-derived risk (OR 1.18, 95% CI 1.08–1.29). EHR-derived prescribing was associated with lower VTE odds (ORs 0.72–0.73).

### Automation feasibility

Model-C achieved an AUROC of 0.704 and AUPRC 0.065, exceeding the expected event-prevalence baseline of 0.023 (Table 4). Incorporating all EHR-derived risk factors in Model-E1 resulted in modest improvements in AUROC (0.718), and AUPRC (0.068) remained largely unchanged. Model-E2, restricted to factors available within 14 hours of admission, showed comparable discrimination (AUROC 0.709) but lower recall (0.409) and higher specificity (0.854). The reduced-feature Model-E3 achieved an AUROC of 0.702 and AUPRC of 0.065. Overall, the EHR-derived models showed only modest differences in discrimination compared with the clinician-recorded baseline. While Model-E1 achieved the highest AUROC, Model-E2 and Model-E3 provided comparable performance using temporally available features. Resampling sensitivity analyses are provided in Supplementary Table S5.

**Table 4.** Performance comparison of machine learning models for VTE risk classification across feature configurations. Performance metrics (95% confidence intervals) are reported for four models trained using different feature sets: Model-C (baseline model using clinician-recorded risk factors), Model-E1 (all EHR-derived risk factors), Model-E2 (EHR-derived risk factors available within 14 hours of admission), and Model-E3 (selected subset of E2 risk factors). Supplementary Table S1 lists the included features. The best-performing classifier for each configuration is shown alongside area under the receiver operating characteristic curve (AUROC), area under the precision-recall curve (AUPRC), recall, and specificity.

| Feature configurations | Classifier | AUROC | AUPRC | Recall | Specificity |
| --- | --- | --- | --- | --- | --- |
| **Model-C** | Random forest | 0.704 (0.676, 0.733) | 0.065 (0.049, 0.089) | **0.655 (0.602, 0.702)** | 0.655 (0.648, 0.663) |
| **Model-E1** | Random forest | **0.718 (0.689, 0.746)** | 0.068 (0.054, 0.094) | 0.533 (0.481, 0.585) | 0.757 (0.751, 0.765) |
| **Model-E2** | XGBoost | 0.709 (0.682, 0.736) | **0.073 (0.053, 0.100)** | 0.409 (0.359, 0.462) | **0.854 (0.848, 0.859)** |
| **Model-E3** | Random forest | 0.702 (0.674, 0.730) | 0.065 (0.049, 0.090) | 0.636 (0.585, 0.687) | 0.659 (0.652, 0.667) |

SHAP analysis was performed on Model-C and Model-E1 to identify the most informative predictors (Figure 6). Across both models, personal or first degree family history of VTE, active cancer or cancer treatment, significant medical morbidities, and age ≥ 60 years old were among the most important predictors for identifying VTE. Notably, the factor ranked most influential in Model-E1, history of VTE, was among those most frequently omitted from clinician-recorded forms (recall 0.05).

**Figure 6.**
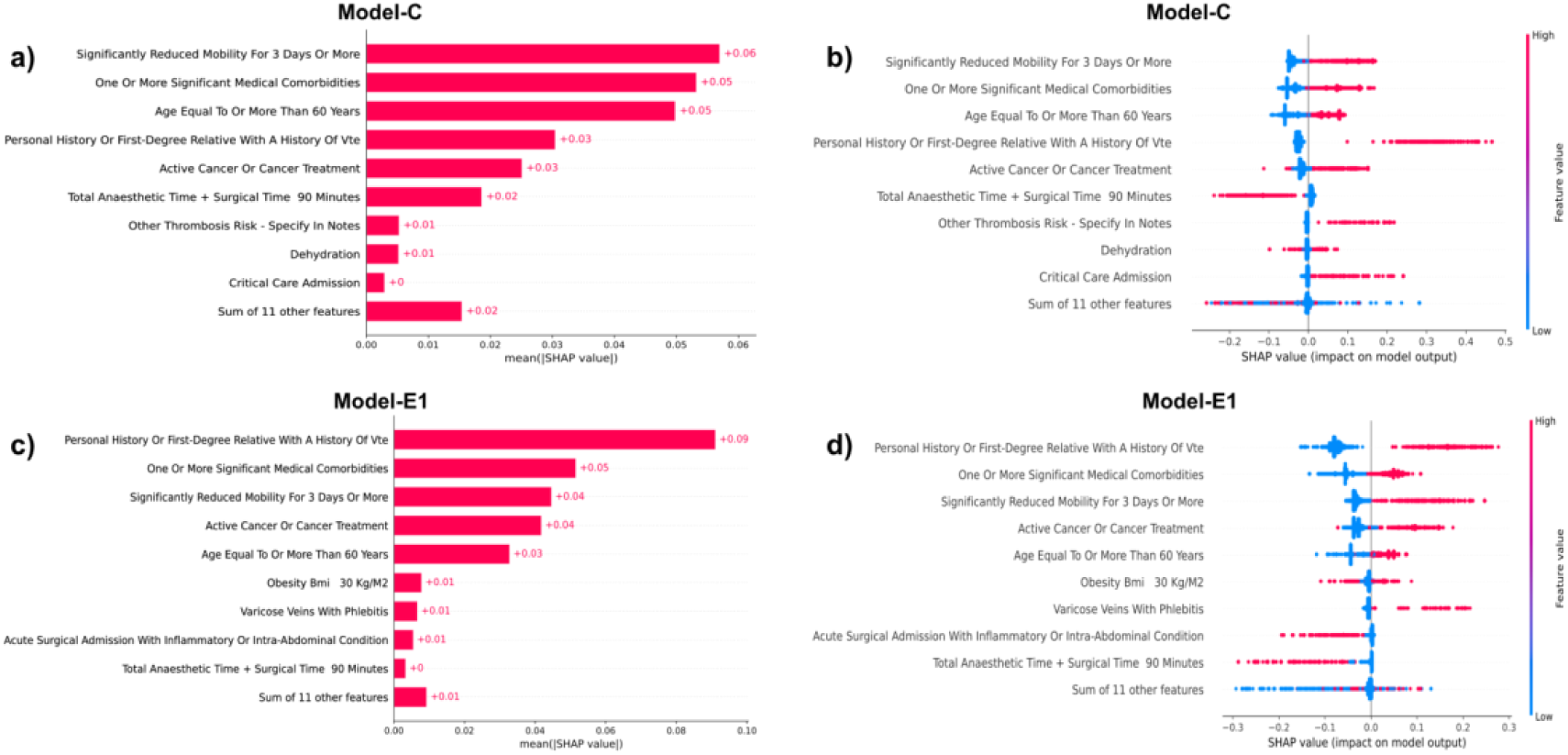
Feature importance and distribution of SHAP values for Model-C and Model-E1 in VTE risk identification. Panels a) and c) show global feature importance ranked by mean absolute SHAP values for Model-C and Model-E1, respectively. Panels b) and d) present SHAP beeswarm plots illustrating the distribution of individual feature values on model output. SHAP values were computed on the held-out test set. Alt text: Four panels show global feature rankings and SHAP-value distributions for clinician-recorded Model-C and EHR-derived Model-E1. VTE history, active cancer or treatment, significant medical comorbidities, and older age are among the most informative features.

## Discussion

This work shows that much of the information required for VTE risk assessment is already present in routinely-collected EHR data. We provide evidence on four related aspects of VTE risk assessment—completion, documentation concordance, clinical validity, and automation feasibility—showing that although structured VTE assessment is embedded in routine clinical practice, important limitations remain. These findings support a role for EHR-based pre-population of risk assessment forms using existing EHR data, reducing reliance on manual data entry while supporting more timely and accurate assessments.

### Completion and documentation quality

Overall completion rates were consistently high, while completion within 14 hours was notably lower, particularly in earlier years. The dip in overall completion around 2020 during the COVID-19 pandemic may reflect clinical priorities, workflow pressures, and evolving thromboprophylaxis guidance [22,23]. Surgical patients were consistently assessed more promptly than medical patients expected to have reduced mobility. This may reflect the more structured nature of surgical pathways, where perioperative processes are often protocolised and time-sensitive, whereas medical admissions may be more heterogeneous and operationally complex. Shifts in inpatient composition, including increases in low-risk medical and day-case-surgical admissions, may also partly explain improvements in observed timeliness.

Despite high completion rates, completion alone is not a reliable indicator of data quality. Discrepancies between clinician-recorded entries and EHR-derived risk factors were common. Agreement varied considerably: structured demographic and major clinical variables were consistently documented, whereas less frequent or more complex risk factors showed poor precision and recall, reflecting a combination of under-recognition, inconsistent documentation practices, ambiguity in clinical interpretation, and limitations in structured extraction. Stronger aggregated performance compared with individual risk factors may reflect pragmatic documentation in a checklist-based system, whereby documenting one or a few thrombosis risk factors is sufficient to signal overall risk. Unlike checklist-based approaches, weighted scoring systems such as Padua or Caprini may be more sensitive to missing variables. Bleeding risk concordance was substantially poorer, likely indicating that bleeding risk factors are less consistently assessed or documented, or that they are more difficult to capture reliably through either form completion or structured EHR definitions.

Similar challenges have been reported in other score-based VTE risk assessment tools [12]. Pre-population directly targets the failure mode we observed, omission of factors that the EHR already contains, while leaving the clinician to confirm, add, or override.

Further discrepancies were observed between documented prophylaxis decisions and prescribing records. Inconsistencies were common in both directions. These findings suggest that risk assessment forms do not fully capture real-world prescribing behaviour, limiting their reliability as a standalone representation of clinical decision-making.

### Clinical validity

Higher VTE odds with thrombosis risk and lower prophylaxis use with bleeding risk support the assessment’s clinical relevance. The documentation gaps resurfaced when estimating the association between prophylaxis and VTE: clinician-recorded prophylaxis showed no protective association, while EHR prescribing data did. The independent association of bleeding risk with increased VTE risk likely reflects patient complexity and pharmacological prophylaxis selection rather than causation. These findings indicate that structured assessment forms contain clinically meaningful information but are limited by incomplete and inconsistent documentation.

### Feasibility of EHR-supported pre-population

The machine learning analyses addressed this question by evaluating whether derived EHR variables contain comparable discriminative information to clinician-recorded risk factors, rather than developing a superior VTE prediction model. EHR-derived features were binarised to match the form.

Models based on EHR-derived features achieved modest improvements over clinician-recorded inputs, indicating that additional useful information exists beyond form data. Importantly, models restricted to variables available within 14 hours performed similarly to full-EHR models, suggesting that much of the useful predictive information is available within the clinically relevant window, and supporting the feasibility of early risk prediction.

Furthermore, reduced-feature models achieved comparable performance, with potential advantages for interpretability and implementation. Although model performance remained constrained by low VTE event prevalence (0.023) and was consistent with previously reported VTE prediction models [24,25], the objective here was feasibility assessment rather than prediction optimisation. SHAP analyses further confirmed that the key risk factors identified by the models were consistent with clinical knowledge, supporting interpretability and clinical plausibility.

Together, these results support the feasibility of partial EHR-assisted pre-population of VTE risk assessment and its potential to close current gaps in documentation, namely incomplete assessments (3.34%) and delayed completion (16.10%), ensuring all eligible patients receive an assessment within the required timeframe. Nevertheless, they highlight the need for further refinement and prospective validation.

### Limitations and future research

This study has several limitations. First, concordance was evaluated against structured EHR-derived definitions, which may themselves be subject to coding errors, incomplete documentation, and clinicians’ subjective interpretation. Second, risk factors and VTE outcomes were primarily defined using structured coding systems such as ICD-10 and SNOMED, which may not capture all clinically relevant information and may lack precise timestamps. Third, free-text reasons for not prescribing prophylaxis were not analysed.

Fourth, the machine learning models did not account for the effects of thromboprophylaxis, which may confound the observed relationship between baseline risk factors and subsequent VTE outcomes. Finally, data came from one NHS Trust and external validation was not performed. Calibration, clinical utility, and subgroup performance require further evaluation before deployment. Future work should validate pre-populated fields and prospectively assess clinician workload. Incorporating clinical notes and radiology reports may improve risk factor and outcome identification.

## Conclusions

The VTE risk assessment pathway was widely completed and demonstrated clinical usefulness. This study identified delayed completion and discrepancies between forms and EHR data, including bleeding risk classification and documented versus prescribed prophylaxis, highlighting opportunities to improve documentation. EHR-derived variables showed discrimination similar to clinician-recorded factors. Pre-populating the mandated form from EHR data offers a pragmatic, scalable route to reduce documentation burden, reduce preventable VTE events, and improve the completeness and timeliness of a long-standing patient safety requirement.

## Acknowledgements

This research was undertaken within the Imperial Secure Data Environment which is run and managed by the iCARE team. iCARE receives funding from the National Institute for Health Research (NIHR) Imperial Biomedical Research Centre (NIHR203323) based at Imperial College Healthcare NHS Trust & Imperial College London. The views expressed are those of the authors and not necessarily those of the NHS, the NIHR, or the Department of Health and Social Care.

## Data Availability

The data analysed during the current study are available from the Imperial Secure Data Environment (SDE), but restrictions apply to the availability of these data and so are not publicly available. The data may be made available to qualified researchers on reasonable request to.

## Code Availability

The underlying code for this study is not publicly available but may be made available to qualified researchers on reasonable request to.

## Author Contributions

Conceptualization: ZL, EY, AR, OWC, MA, TS, KG, BG, EM. Data curation: ZL, AR, OWC, MA, TS. Formal analysis: ZL, OWC, MA, TS. Funding acquisition: EM. Investigation: ZL, EY, AR, OWC, MA, TS, KG, CB, ZK, GH, EM. Methodology: ZL, EY, AR, OWC, MA, TS, KG. Project administration: EY, BG. Resources: BG. Software: ZL, OWC. Supervision: KF, EM. Validation: ZL, EY, OWC, MA, TS, KG, CB, ZK, GH, BG, EM. Visualization: ZL, MA. Writing -original draft: ZL, EY, KF, EM. Writing - review and editing: all authors.

## Supporting information

Supplementary eligibility criteria, electronic health record extraction definitions, analysis sample sizes, model sensitivity analyses, and agreement matrices. Includes Notes S1-S2, Tables S1-S5, and Figure S1.

